# COVID-19 among nursing staff: Settings and regional differences

**DOI:** 10.1101/2020.08.14.20174797

**Authors:** Manuela Hoedl, Silvia Bauer, Doris Eglseer

## Abstract

**Purpose:** This study was carried out to describe settings of and regional differences in the occurrence of COVID-19 among nursing staff, analysing COVID-19 symptoms, testing and diagnosis procedures.

**Design:** We used an online survey to conduct this cross-sectional study among nursing staff in different settings. Data collection was carried out between 12 May and 13 July 2020.

**Methods:** The survey included questions that allowed us to collect demographic data (e.g. age), professional qualifications (e.g. nurse, nurse aid) and data regarding the federal states and settings in which the participants worked. In additon, we asked the participants to describe their COVID-19 symptoms, if any were experienced, and the result of COVID-19 testing that they underwent. We used descriptive statistics as well as bivariate analysis methods to calculate differences.

**Findings:** More than 80% of the nursing staff (N = 2600) were women, nearly half of these staff members worked in the province of Styria and about three-quarters worked in hospitals. In general, nearly every sixth nurse reported experiencing COVID-19 symptoms. We found statistically significant differences between the settings and the federal provinces with regard to the COVID-19 symptoms reported, but not the test results. The highest porportion of nurses who experienced symptoms worked in lower Austria and in the primary care sector. In total, 1.6% of the participating nurses were tested for COVID-19, as well as about 1% of the nurses who worked in the nursing homes. The highest number of tests were carried out in Vienna followed by the province of Burgenland. In total, all of the nurses who underwent testing were diagnosed with COVID-19.

**Conclusions:** Only 1% of the nurses who worked in nursing homes were tested. This group of nurses takes care of the most vulnerable and high-risk group in Austrian society.

Therefore, the nursing home staff should have the possibility to be tested even though they did not experience/report any symptoms. The highest prorportion of nurses who experienced symptoms worked in the primary care sector. In the future during a pandemic, health care staff should be comprehensively tested in all settings.

**Clinical Relevance:** Providing insights into COVID-19 symptoms experienced by nursing staff workforce and testing for COVID-19 can help us address these aspects in future pandemics more efficiently. In addition, these insights can help to shift the perspectives from critical and acute settings to other settings, e.g. nursing homes. This is an important change in perspectives, as these nurses have worked with the most vulnerable and high-risk group during the COVID-19 pandemic. Ensuring the safety of nursing home residents and providing, high-quality nursing care and interventions can reduce hospital admission rates, decrease health care costs during such pandemics and might even reduce secondary morbidity and mortality.

## Introduction

On 13 August 2020 the World Health Organization reported more than 20 million confirmed cases of the Coronavirus disease (COVID-19) worldwide, including more than 510,000 deaths (WHO, 2020). These numbers reflect the devasting pressure on and challenges in the health care systems worldwide.

International databases are being updated daily in order to measure the incidence of the severe acute respiratory syndrome coronavirus 2 (SARS-CoV-2) as well as track the number of persons diagnosed with COVID-19 over time. These databases include the: (1) the WHO Coronavirus Disease Dashboard (WHO, 2020), (2) U.S. Centers for Disease Control and Prevention (U.S. Centers for Disease Control and Prevention, 2020a), (3) EU Open Data Portal of the European Centre for Disease Prevention and Control (European Centre for Disease Prevention and Control, 2020), or even national dashboards such as (4) the Official COVID-19 Dashboard for Austria (Federal Ministry of Social Affairs, 2020) or the COVID-19 Dashboard maintained by the Robert Koch Institut in Germany (Robert Koch Institute, 2020). However, all of these databases are population-based databases and do not place a focus on high-risk groups such as health care workers. With regard to COVID-19, health care workers are recognised as a specific high-risk group (U.S. Centers for Disease Control and Prevention, 2020b).

Nurses and midwives make up the largest group (i.e. about 50%) of all health care workers, and (WHO, 2006). Especially nurses have been recognised as key players in crisis and postcrisis situations (WHO, 2016), such as the COVID-19 pandemic.

In recent months, hundreds of papers have been published internationally on the topic of COVID-19. Many of the studies describe vaccine development efforts, specific treatment interventions (Ahmed, Quadeer, & McKay, 2020; Batlle, Wysocki, & Satchell, 2020; Garcia Filho, 2020; Li et al., 2020; Liu, Zheng, & Wang, 2020), or the pathophysiology of the underlying virus (Forster, Forster, Renfrew, & Forster, 2020; Gildenhuys, 2020; He, Tao, Yan, Huang, & Xiao, 2020; Jaimes, André, Chappie, Millet, & Whittaker, 2020; Kiyotani, Toyoshima, Nemoto, & Nakamura, 2020; Lan et al., 2020; Lau et al., 2020).

Some of the papers place a focus on epidemiology (Corbett et al., 2020) or lessons learned during COVID-19 (Albalate et al., 2020; D’Adamo, Yoshikawa, & Ouslander, 2020; Kim et al., 2020; Sacco, Foucault, Briere, & Annweiler, 2020). Others have been carried out to investigate how COVID-19 has affected patients (Lin, Lu, Cao, & Li, 2020; Mokrzycki & Coco, 2020; Nowak et al., 2020), nursing home residents (Kimball et al., 2020), or older persons in the community (Nikolich-Zugich et al., 2020).

We conducted a systematic literature review of these papers and found only a few studies that specifically addressed nursing staff and COVID-19 (Bird, Badhwar, Fallon, Kwok, & Tang, 2020; Guery et al., 2020; Halcomb et al.; Millar, 2020; Mo et al., 2020; Ning et al., 2020; Yifan et al., 2020; H.-j. Zhang et al., 2020; Y.-P. Zhang et al., 2020). Some of these papers described the consequences of COVID-19 in relation to nursing staff (Mo et al., 2020; Ning et al., 2020; H.-j. Zhang et al., 2020; Y.-P. Zhang et al., 2020). One study was carried out on nurses as COVID-19 patients (Millar, 2020); one study, on a symptom cluster of ICU nurses who treated COVID-19-positive pneumonia patients (Yifan et al., 2020); and one study, on SARS-CoV-2 infection rates among respiratory staff nurses (Bird et al., 2020). In another recent study, researchers investigated the experiences of nurses who worked in the Australian primary health care sector during the COVID-19 pandemic (Halcomb et al.).

However, none of the above-mentioned publications provided detailed insights into the nursing staff’s COVID-19 symptoms, testing procedures/results, or diagnoses. In addition, based on the results of the literature review, no study has been thus far carried out to investigate the effects of settings and regional differences on reported symptoms and testing. However, such a study could provide researchers with important, comprehensive insights into COVID-19 infection rates in nursing staff. Furthermore, these study findings could indicate the settings and regions in which testing should start earlier if another pandemicoccurs. Hence, we designed and carried out the current study to investigate settings and specific regional differences with regard to COVID-19 symptoms, testing and diagnoses among nursing staff.

## Methods

### Design, settings and sample

This study was conducted as a cross-sectional study, using an online questionaire. In order to reach our target sample, we applied the snowball sampling strategy and used social media. We developed a questionaire that was subsequently distributed to nursing staff throughout Austria, independent of their qualifications (e.g. nurse, nurse aid) and work settings (e.g. hospital, nursing homes). The main target group included nurses who were working at the bedside during the COVID-19 pandemic in Austria.

### Data collection instrument and analysis

We collected data from 12 May to 13 July 2020. For this study, we collected the following data: (1) demographics (e.g. age, gender) (2) region, (3) setting, (4) professional qualification (e.g. nurse, nurse aid), (5) job experience in years, (6) symptoms of COVID-19 experienced (Yes/No) and (7) if the participants had been tested positive for COVID-19 (Y es, I was tested positive for COVID-19/No, I was tested negative for COVID-19/ No, I have not been tested for COVID-19 at all).

The main variables in this study were the region, setting and if the nurses had been tested, including the results. All of these variables were categorical variables for which we calculated descriptive statistics in percentages. We also conducted a bivariate analysis using the X^2^ test and Cramér’s V as the effect size. A p-value of <.05 indicates a statistically significant result in this study.

### Ethics

On the first page of the online questionnaire, we provide the participants with information about the aim of the study, contact persons as well as about data security. All participants then had to sign a written informed consent for, in compliance with the General Data Protection Regulation of the European Union, in order to participate and continue to the questionnaire. All data were collected anonymously. An ethical approval was granted by the responsible ethical committee.

## Findings

In total, 3497 nursing staff members participates in the study, resulting in a large sample of questionnaires filled out online. Of this number, 892 questionnaires (25.5%) were incomplete: 56 of the invited persons chose not to participate; 567 participated, but stopped did not complete the questionnaire; and 269 only read the welcome page of the questionnaire. In addition, we had to exclude data from five participants, as they were 65 years or older, which is the retirement age in Austria. As a result, we were able to include data from 2600 persons in this analysis. Table 1 depicts the sample characteristics.

**Table 1.** Sample characteristics

|  | **Nursing staff (N = 2600)** |
| --- | --- |
| Female % (n) | 83.7 (2177) |
| Mean age in years (SD) | 38.1 (11.0) |
| Settings % (n) |  |
| Home care | 2.7 (70) |
| Home for disabled persons | 2.1 (54) |
| Hospital | 73.3 (1905) |
| Nursing home | 17.2 (448) |
| Others | 2.8 (73) |
| Primary care | 0.4 (11) |
| Rehabilitation | 1.5 (39) |
| Federal province where nurses worked % (n) |  |
| Burgenland | 8.7 (227) |
| Carinthia | 3.9 (101) |
| Lower Austria | 6.8 (177) |
| Salzburg | 7.7 (201) |
| Styria | 49.5 (1287) |
| Tyrol | 8.1 (210) |
| Upper Austria | 6.5 (170) |
| Vienna | 5.3 (137) |
| Vorarlberg | 3.5 (90) |

More than 80% of the nurses were women, and nearly half of the nurses worked in the province of Styria (49.5%), Burgenland (8.7%), or Tyrol (8.1%). Nearly three-quarters of the nurses worked in a hospital setting (73.3%). Table 2 describes the setting and regional differences with regard to COVID-19 symptoms and testing reported by nursing staff.Table 2. Settings and regional differences with regard to symptoms of and testing for COVID-19 reported by nursing staff.

**Table 2.** Settings and regional differences with regard to symptoms of and testing for COVID-19 reported by nursing staff

|  | Nursing staff |  |
| --- | --- | --- |
|  | With symptoms* | Tested |
| Austria (*N* = 2600) | 15.0 (389) | 1.6 (41) |
| Settings % |  |  |
| Home care (*n* = 70) | 12.9 (9) | 1.4 (1) |
| Home for disabled persons (*n* = 54) | 13.0 (7) | - |
| Hospital (*n* = 1905) | 16.3 (310) | 1.8 (35) |
| Nursing home (*n* = 448) | 9.8 (44) | 1.1 (5) |
| Others (*n* = 73) | 13.7 (10) | - |
| Primary care (*n* = 11) | 36.4 (4) | - |
| Rehabilitation (*n* = 39) | 12.8 (5) | - |
| Federal province where nurses work |  |  |
| Burgenland (*n* = 227) | 17.6 (40) | 2.6 (6) |
| Carinthia (*n* = 101) | 10.9 (11) | - |
| Lower Austria (*n* = 177) | 22.6 (40) | 1.7 (3) |
| Salzburg (*n* = 201) | 16.9 (34) | 1.5 (3) |
| Styria (*n* = 1287) | 12.9 (166) | 1.7 (22) |
| Tyrol (*n* = 210) | 17.1 (36) | 1.4 (3) |
| Upper Austria (*n* = 170) | 17.6 (30) | - |
| Vienna (*n* = 137) | 16.8 (23) | 2.9 (4) |
| Vorarlberg (*n* = 90) | 10.0 (9) | - |

\*Statistically significant difference between the settings, $V = .080$ , $p = .013$ ; Statistically
significant difference between the provinces, $V = .086$ , $p = .011$ ;

Nearly every sixth nurse (15%) reported experiencing COVID-19 symptoms. Of these nursing staff members, 16.3% of the staff in hospitals and 9.8% of the staff in nursing homes experienced COVID-19 symptoms. More than one-third of the nurses who worked in the primary care sector reported experiencing COVID-19 symptoms. We found a statistically significant difference with regard to the regional setting and COVID-19 symptoms. The highest prorportion of nurses who reported symptoms worked in lower Austria (22.6%), followed by Upper Austria and Carinthia (17.6%). In contrast, 10% of the nurses who worked in Voralberg reported that they experienced COVID-19 symptoms.

Our findings show that 1.6% of the participating nurses were tested for COVID-19. We could not identify any statistically significant difference between the settings (V =.041, *p* =.575)as well as the federal provinces (V =.060, *p* =.309), with regard to COVID-19 testing. The federal provinces where the most tests for nursing staff were conducted were Vienna (nearly 3%) followed by Burgenland. All of the tested nurses (n = 41), independent of the federal province or setting in which they worked, were diagnosed with COVID-19.

## Discussion

In this study, we investigated the effect of settings and regional differences on the reporting of COVID-19 symptoms, testing and diagnosis by nursing staff.

In general, nearly every sixth participating nurse (N= 2600) reported experiencing COVID-19 symptoms. We found statistically significant differences between the settings and the federal provinces with regard to the COVID-19 symptoms experienced. The highest prorportion of nurses who experienced symptoms worked in lower Austria, followed by Upper Austria and Carinthia. Nearly 10% of nursing home staff and about 16% of staff working in the hospitals reported experiencing COVID-19 symptoms.

We identified one study in which the researchers focused on symptoms in COVID-19-positive health care workers (Bird et al., 2020). The authors described that most of the COVID-19-positive staff experienced at least one systemic symptom (i.e. any combination of the following: fever, headache, myalgia, or fatigue; *n* = 37, 78.7%) and at least one respiratory symptom (i.e. any combination of the following: cough, sore throat, shortness of breath, or chest tightness/pain, *n* = 41, 87.2%). In the health care workers who received a negative PCR test result, 59.4% (n = 95) reported at least one systemic symptom, and 71.9% (n = 115) reported at least one respiratory symptom. The differences between this study’s results and our results could be explained by the fact that we included not only nursing staff that had been tested. In Austria, the Ministry for Social Affairs, Health, Care and Consumer Protection issued instructions for health care workers, specifically instructing them to perform personal observations and self-assessment of their symptoms. If they experienced a cough or fever, they were requested to call an established ‘health advice hotline’ number (1450) to receive more information. The person contacted via this number would then make the decision regarding whether someone would be tested or not.

Some of our results were surprising; we did not expect the highest prorportion of nurses who experienced symptoms to have worked in lower Austria, followed by Upper Austria and Carinthia. These results were surprising, because the first regions to report a high prevalence of COVID-19 were Tyrol (Ischgl, Kappl, See, Galtür und St. Anton am Arlberg) and Salzburg (Pongau) (Nagl, 26. March 2020). However, these findings might be explained by the fact that we asked the nursing staff retrospectively; at the time of our data collection, the provinces of Lower and Upper Austria were reporting relatively high numbers of COVID-19-affected persons.

Although we could not identify any statistically significant difference between the federal provinces or settings with regard to COVID-19 testing, we believe that it is of great importance to point out that the federal provinces with the highest numbers of tests were Vienna followed by Burgenland. In general, from the beginning of the COVID-19 pandemic in Austria up until 3 August, 910,437 persons were tested. Of this number, 25% were from Vienna, 17.7% were from Tyrol and 17.4% were from Lower Austria (Federal Ministry of Social Affairs, 2020). These data contrast with our results. This might be explained by the fact that we only asked nurses if they had been tested, and the data for the above-mentioned tests includes all tests conducted in Austria during this period. However, we find this contrast surprising, as the federal province of Burgenland had very low rates of COVID-19-affected persons as compared to the other federal provinces, such as Tyrol. One explanation for these findings might be the short distance between Vienna and Burgenland, i.e. the testing teams from Vienna might also have conducted tests in Burgenland.

In total, all of the tested nurses (n = 41), regardless of their settings or the federal province, were diagnosed with COVID-19. In the study of Bird et al. (2020), 63 nurses participated in the study and 34.9% of these tested positive (Bird et al., 2020). Two other studies reported a similar percentage of COVID-19-positive health care workers. In these studies, 17% (Chen et al., 2020) and 29.1% (Tang et al., 2020) of the nurses tested positive for COVID-19. The difference between the results reported in these studies and our results might be explained by the relatively small number of nurses tested in general.

However, in our study, we showed that all the nurses tested were diagnosed with COVID-19. This had two major consequences. First, if more nurses had been tested, we could assume that Austria would have a higher COVID-19 incidence. Second, considering that many persons are asymptomatic, and nurses who do not experience symptoms are not tested, these nurses might become “superspreaders”. This aspect should be carefully considered now when normal life is starting again.

### Limitations

Every study has strengths and limitations. The first limitation of our study is that our sample is not representative for the Austrian health care setting. As an example, according to data collected in 2017 (Federal Ministry of Labor, 2019), 53% percent of all nurses were employed at hospitals, 33% in nursing homes and 14% in home care settings. In our study, nearly three-quarters of the nurses who participated worked in a hospital setting, only 17% worked in nursing homes and 2.7% worked in the home care setting. Another limitation of this study might be that we asked the nurses retrospectively regarding their experiences with COVID-19. The COVID-19 pandemic began in Austria at the beginning of March, with a lockdown order issued on 16 March 2020. When we started the survey (May 12), shopping centers, shops, restaurants, cafes, bars and nursing homes had already opened again. This retrospective view might have introduced a bias. However, most of the participants completed the questionnaire within the first two weeks after the online platform was opened.

## Conclusions

In general, only 1.1% nurses in nursing homes were tested for COVID-19. Nurses who are taking care of the most vulnerable and high-risk group of patients should be given the possibility to be tested more frequently for COVID-19, even if they do not experience any symptoms. This aspect is of particular importance, as nurses are likely to spend more time with the patients/residents than any other health care professional and have been recognised as key players in the COVID-19 crisis. The highest prorportion of nurses who experienced symptoms were working in Lower Austria at the time of the survey, and most of the tests were performed in a hospital setting. In the future, testing in all settings is highly recommended in a pandemic. We could not identify any statistically significant difference between the federal provinces or the settings with regard to the COVID-19 testing. However, most of the tests were made in Vienna followed by Burgenland, even though Tyrol initially had the highest numbers of affected persons. In the future, testing performed in a pandemic should be based on the rates of suspected and affected cases, starting with the primary care sector.

## Data Availability

Due to legal issues, data can not be made available.

